# Effects of Morning Bright Light Therapy on Sleep, Alertness, Mood, and Cognition in Healthy University Students: A Randomized Crossover Trial

**DOI:** 10.64898/2026.07.04.26357282

**Authors:** Cehao Yu, Caicai Zhang, Hayley Tsang, Lu Li, Nayantara Santhi

## Abstract

**Objectives:** To test whether one week of self-administered morning bright light therapy (BLT) improves sleep, daytime sleepiness and alertness, mood, and objective cognition in healthy university students.

**Methods:** Thirty-three healthy students completed a two-week randomized within-subject crossover trial comparing one week of morning BLT (30 min of 10,000 lx; melanopic equivalent daylight illuminance ≈ 8,989 lx) with one week of usual-light control in counterbalanced order, with no washout. Sleep was assessed with wrist-worn Fitbit sleep tracking and daily diaries; daytime sleepiness (Karolinska and Stanford Sleepiness Scales), positive and negative affect (PANAS), mood (POMS), and a cognitive battery (Stroop, Flanker, Corsi, verbal span) were also assessed, alongside post-trial semi-structured interviews. Outcomes were analyzed with linear mixed-effects models, with Holm correction across five primary outcomes.

**Results:** BLT reduced daytime sleepiness in a time-of-day–specific manner (condition × time-of-day interaction; largest reduction at 12:00, dz = −0.58, with a smaller but still significant reduction at 15:00), reduced night-to-night variability in sleep duration (dz = −0.52), increased Fitbit sleep efficiency (dz = 0.81), and increased PANAS positive affect (dz = 0.41). Objective cognition was unchanged across all measures. Interviews indicated that participants experienced BLT primarily as a sleep and alertness intervention, with minor tolerability issues.

**Conclusions:** Brief morning BLT improved alertness, sleep regularity and efficiency, and positive affect, but not objective cognition, in healthy students, supporting morning light as a low-burden strategy for daytime functioning while cautioning against overstating cognitive benefits.

## Introduction

Adequate, well-timed sleep is critical for daytime alertness, mood, and learning. However, insufficient and irregular sleep remains widespread among university students and carries measurable costs for daytime functioning and academic performance (Hershner & Chervin, 2014; Phillips et al., 2017). One potentially modifiable contributor to poor sleep in this population is daily light exposure. Contemporary indoor environments often produce a pattern we term ‘light inversion’, an attenuation of natural light–dark patterns, characterised by dim daytime light and excessive evening light. In particular, evening exposure to artificial light suppresses melatonin secretion and delays circadian phase, resulting in later sleep onset and reduced alertness the following morning (Brown et al., 2022; Chang et al., 2015; Santhi et al., 2012). When combined with inadequate daytime light, this pattern can lead to misalignment between endogenous circadian time and externally imposed social schedules, the so-called social jetlag (Roenneberg et al., 2012). It has been linked to daytime fatigue and impairments in mood and cognition (Wittmann et al., 2006).

Bright light therapy (BLT), typically administered ∼10,000 lx for 30 minutes in the morning, is an established non-pharmacological treatment for seasonal and non-seasonal depression, with efficacy comparable to antidepressant medication (Golden et al., 2005; Lam et al., 2016). Beyond clinical populations, BLT has been shown to acutely enhance subjective alertness and vigilance compared with dim light exposure (Cajochen, 2007; Phipps-Nelson et al., 2003). These effects are mediated via a non-image-forming (NIF) eye-to-brain pathway driven by melanopsin-containing intrinsically photosensitive retinal ganglion cells. These ganglion cells project to the suprachiasmatic nucleus (SCN), enabling light to entrain circadian rhythms (Czeisler et al., 1999) and to acutely suppress melatonin and enhance alertness (Berson et al., 2002; Lucas et al., 2014). The SCN relies on consistent daily light input to maintain alignment with the 24-hour day (Czeisler et al., 1999). In interaction with homeostatic sleep pressure, it regulates sleep–wake dynamics and daytime performance (Borbély et al., 2016; Dijk & Czeisler, 1995), providing a plausible pathway through which BLT may influence sleep, mood, and cognition. Accordingly, melanopic measures are increasingly used to quantify light exposure beyond conventional photopic lux (Brown et al., 2022; Lucas et al., 2014). Nevertheless, the evidence for NIF effects on sleep and especially cognition remains mixed. Acute alerting responses vary substantially across experimental contexts (Souman et al., 2018), and findings for objective cognitive performance in healthy individuals are inconsistent, with many studies reporting null effects (Vandewalle et al., 2009).

Several important gaps in the literature remain. First, much of the BLT literature derives from clinical populations or tightly controlled laboratory settings, with limited examination of ecologically valid, self-administered use in healthy young adults. Second, circadian disruption in university students is more often driven by lifestyle factors, such as late-night screen use, academic demands, irregular schedules, and predominantly indoor living, rather than seasonal variation in photoperiod (Hershner & Chervin, 2014). This issue is particularly salient for low-latitude urban environments such as Hong Kong, where seasonal changes in photoperiod are modest. Although outdoor daylight is abundant, it does not translate to the indoor setting since buildings in these regions are designed to balance daylight access with solar heat gain, glare, and air-conditioning demands (Huang et al., 2014).

Against this background, the present study employed a two-week randomized within-subject crossover design, in healthy university students in Hong Kong, contrasting one week of morning BLT (30 minutes, 10,000 lx, melanopically characterized) with habitual-light exposure. Sleep was assessed via wrist-worn actigraphy (Fitbit) and daily diaries, alongside measurements of daytime sleepiness, alertness, mood, and objective cognition, and interviews to assess BLT acceptability. We hypothesized that BLT would reduce daytime sleepiness, improve sleep regularity, and enhance positive affect, with no directional prediction for cognitive outcomes.

## Methods

### Participants

Thirty-three healthy university students from The Hong Kong Polytechnic University were enrolled. Participants were recruited through campus flyers and mailing lists and were required to reside in the Ho Man Tin student halls for the duration of the study. Eligibility criteria were no history of insomnia or other diagnosed sleep disorder, no shift work in the preceding six months, and no use of medications known to affect sleep or alertness. All participants were non-smokers, reported no regular alcohol use, and agreed to abstain from alcohol throughout the two-week protocol. At screening, obstructive sleep apnoea risk was assessed with the Berlin Questionnaire (Netzer et al., 1999) and sleep quality with the Pittsburgh Sleep Quality Index (PSQI; Buysse et al., 1989); individuals with poor sleep quality (global PSQI score > 5) were excluded. Chronotype was assessed with the Morningness–Eveningness Questionnaire (MEQ; Horne & Östberg, 1976), and body mass index was recorded at intake.

Baseline participant characteristics are summarised in Table 1.

**Table 1.** Baseline Participant Characteristics.

| Characteristic | Value |
| --- | --- |
| N | 33 |
| Treatment sequence — BLT first / Control first, n | 17 / 16 |
| Age, years — M (SD) [range] | 24.7 (2.4) [18.0–30.0] |
| Gender — Female, n (%) | 24 (72.7%) |
| Gender — Male, n (%) | 9 (27.3%) |
| BMI, kg/m<sup>2</sup> — M (SD) [range] | 21.1 (2.8) [16.6–28.1] |
| MEQ total score — M (SD) [range] | 42.1 (8.1) [32.0–60.0] |
| MEQ: Definite morning, n (%) | 0 (0.0%) |
| MEQ: Moderate morning, n (%) | 2 (6.1%) |
| MEQ: Intermediate, n (%) | 13 (39.4%) |
| MEQ: Moderate evening, n (%) | 18 (54.5%) |
| MEQ: Definite evening, n (%) | 0 (0.0%) |
| Time in bed — Control week, min, M (SD) | 501.9 (55.7) |
| Wake time — Control week, min past midnight, M (SD) | 564.8 (66.8) |
| KSS at 12:00 — Control week, M (SD) | 4.4 (1.6) |

*Note.* Values are mean (SD), mean (SD) [range], or n (%) unless otherwise indicated. BMI =
body mass index; BLT = bright light therapy; KSS = Karolinska Sleepiness Scale; MEQ =
Morningness–Eveningness Questionnaire.

The study protocol was approved by the Institutional Review Board of The Hong Kong Polytechnic University (reference number HSEARS20240430005) and was conducted in accordance with the Declaration of Helsinki. All participants provided written informed consent before enrolment.

Fitbit-derived analyses were restricted to participants with usable Fitbit data; all participants contributed to questionnaire-, diary-, interview-, and cognitive-based outcomes.

### Study design

The study used a randomised, two-period, two-condition crossover design (Figure 1).

**Figure 1.**
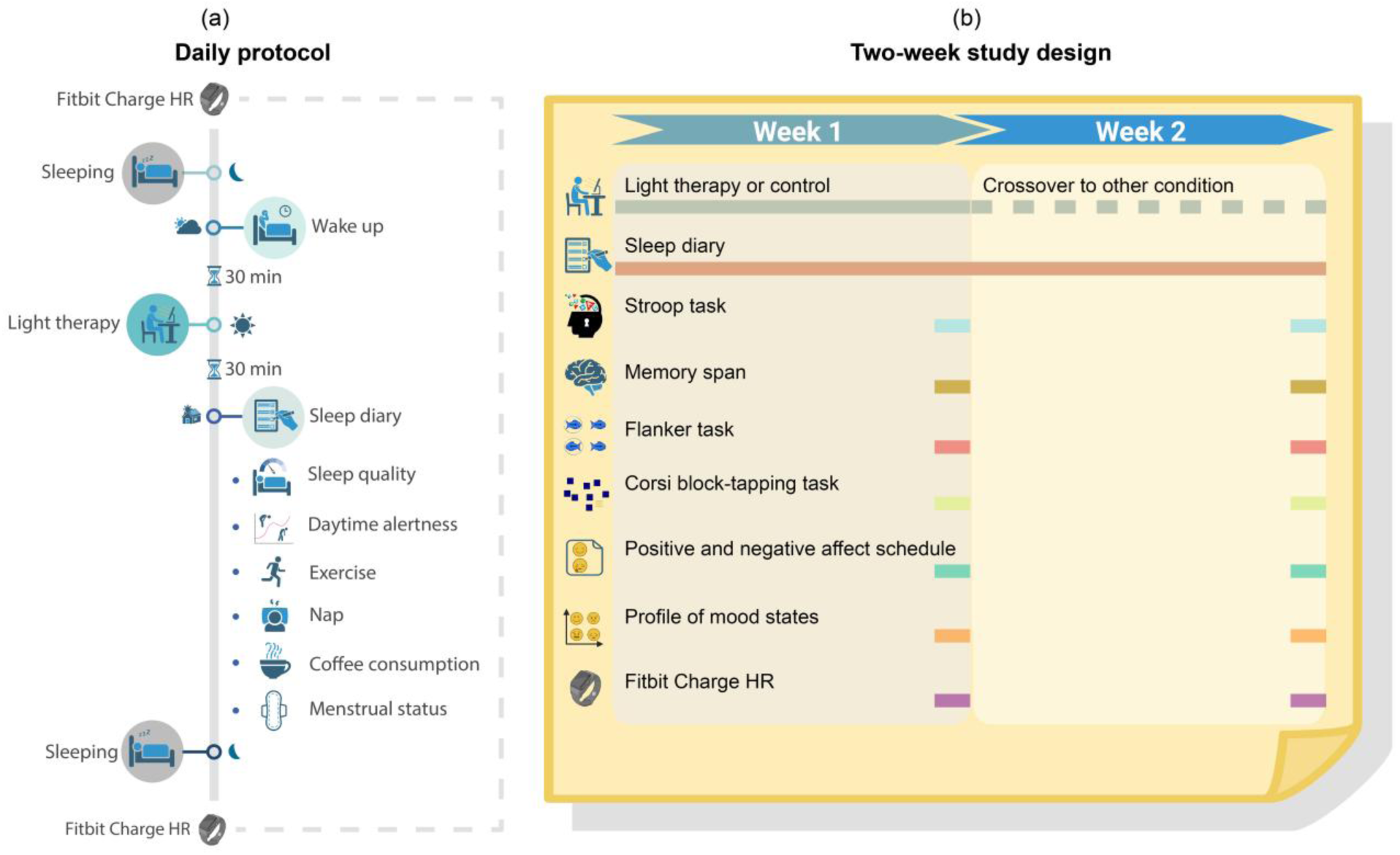
Study Design and Daily Protocol. *Note.* (a) Daily protocol: participants wore a wrist-worn Fitbit Charge HR continuously; on intervention days they completed a 30-min bright light therapy (BLT) session approximately 30 min after waking; and they completed a daily diary covering sleep and daytime items (alertness [KSS, SSS], naps, exercise, caffeine, and—for female participants—menstrual status). (b) Two-week randomized within-subject crossover: one week of BLT and one week of usual-light control in counterbalanced order, with no washout. The daily diary was completed every day and Fitbit monitoring covered the entire study; the cognitive battery (Stroop, Corsi block-tapping, Flanker, and verbal memory span) and the mood scales (PANAS, POMS) were administered on the final day of each condition week. BLT = bright light therapy; KSS = Karolinska Sleepiness Scale; PANAS = Positive and Negative Affect Schedule; POMS = Profile of Mood States; SSS = Stanford Sleepiness Scale.

Each participant completed one week of morning bright light therapy (BLT) and one week of a control condition in counterbalanced order. No washout period was included; the two weeks were consecutive. Period and sequence effects were therefore modelled in all crossover analyses, and carryover was evaluated in sensitivity analyses.

Participants were allocated to one of two treatment sequences by computer-generated block randomisation: BLT in Week 1 followed by control in Week 2, or control in Week 1 followed by BLT in Week 2. No separate allocation-concealment or blinding procedures were documented; the study was therefore conducted as an open-label randomised crossover trial.

### Intervention and control condition

#### Bright light therapy condition

During the BLT week, participants self-administered a 30-minute bright-light exposure session each morning for seven consecutive days, beginning approximately 30 minutes after waking. They were seated approximately 17 cm from the lamp with their gaze generally oriented toward the light source; quiet reading or device use was permitted during exposure.

BLT was delivered with a diffuse, disk-shaped LED light box (Dorui GLD-1005; Guangdong, China; diameter 24 cm). The device offered three factory correlated-colour-temperature (CCT) presets (approximately 2800–3000 K, 4000–4200 K, and 5500–5700 K) and three intensity levels; the high-CCT, high-intensity preset was used. At the experimental geometry (17 cm from the eyes, with the lamp centre slightly below eye level), the spectral power distribution (SPD) and corneal vertical illuminance were measured with a Konica Minolta CL-500A illuminance spectrophotometer. At this setting the device delivered 10,000 lx photopic illuminance, CCT 6010 K, Duv +0.0078, TM-30 Rf 89, and TM-30 Rg 97. From the measured SPD, melanopic equivalent daylight illuminance (melanopic EDI; CIE S 026; International Commission on Illumination, 2018) was 8989 lx (melanopic-to-photopic ratio 0.90), corresponding to an approximate melanopic dose of 4495 lx·h per 30-minute session.

#### Control condition

During the control week, participants used no light box and followed their usual morning routine under ambient room lighting, with no prescribed morning bright-light exposure. Because the control condition included no sham or dim-light device, the study should be interpreted as a randomised crossover comparison of BLT with a usual-light control rather than as a placebo-controlled trial.

### Adherence and tolerability

BLT session completion was recorded in the daily diaries, and continuous Fitbit wear and daily sleepiness ratings were used to monitor participation and outcome completion. Because the protocol prescribed a fixed dose, dose–response analyses based on between-person variation in delivered exposure were not conducted.

Adverse events were monitored throughout the trial through daily contact with participants—maintained by three resident researchers (authors) who lived in the same student-hall block—and through the daily diary, in which participants recorded any adverse effects; a structured adverse-event checklist was not administered.

### Outcome measures

#### Primary outcomes

The primary confirmatory outcome family was defined a priori in the approved study protocol, on the basis of the study’s hypothesized domains rather than observed effects. For each domain in which morning bright light was hypothesized to act—sleep, daytime sleepiness, affect, and cognition—a single, theoretically representative index was designated as primary, with processing speed and working memory treated as two distinct cognitive constructs. These five outcomes constituted the primary confirmatory family and were corrected for multiple comparisons with the Holm sequential Bonferroni procedure:

- Sleep regularity: the within-person standard deviation of nightly sleep duration across each condition week, derived from Fitbit data (Fitbit-analysis subsample).
- Daytime sleepiness: the condition × time-of-day interaction in Karolinska Sleepiness Scale (KSS; Åkerstedt & Gillberg, 1990) ratings, assessed repeatedly across the day in the daily diary.
- Positive affect: the Positive and Negative Affect Schedule (PANAS; Watson et al., 1988) Positive Affect score, administered on the final day of each condition week.
- Processing speed: log-transformed reaction time on congruent trials of the Stroop task.
- Working memory: the mean verbal span score across the digit, letter, and word span subtests.

#### Secondary and exploratory outcomes

All remaining measures within these domains were specified in the protocol as secondary and interpreted as supporting or exploratory. Secondary sleep outcomes included Fitbit-derived sleep efficiency, total sleep duration, time in bed, sleep onset latency, wake time, and sleep midpoint; diary-derived time in bed and wake time; and Stanford Sleepiness Scale (SSS; Hoddes et al., 1973) ratings on the same daily schedule as the KSS. Secondary mood outcomes included PANAS Negative Affect and Profile of Mood States (POMS; McNair et al., 1971) subscales, including Total Mood Disturbance, Anger–Hostility, and Vigor–Activity, together with a POMS-derived Esteem-related Affect–Positive composite comprising the items proud, competent, confident, and satisfied. Secondary cognitive outcomes included Stroop reaction time (ms) and Stroop interference, Flanker congruent reaction time and interference, and Corsi block-tapping forward and backward span. Circadian-characterisation outcomes included the alignment between MEQ-predicted and observed sleep midpoint, and weekday–weekend social jetlag.

### Assessment schedule and procedure

Participants attended a baseline briefing session at which they received written and verbal instructions. They were asked to limit caffeine intake to no more than two cups of coffee (or equivalent) per day and to record their intake in the daily diary. Each participant then completed two consecutive weeks of monitoring—one BLT week and one control week—in randomised order.

Each day, participants wore a Fitbit Charge HR continuously and completed a daily diary. The diary comprised sleep items and daytime items; the latter included KSS and SSS sleepiness ratings, naps, exercise, caffeine consumption, and—for female participants—menstrual status. KSS and SSS ratings were recorded at 12:00, 15:00, 18:00, 21:00, and before bed. The primary diurnal-profile analyses used the four daytime assessments from 12:00 to 21:00; bedtime ratings were retained as diary variables but were not entered into the primary linear time-of-day interaction model. On BLT days, the 30-minute session was completed approximately 30 minutes after waking.

The cognitive battery and the PANAS were administered on the final day of each condition week. Cognitive tasks comprised the Stroop, Flanker, and Corsi block-tapping tasks and verbal memory span tasks for digits, letters, and words. The POMS was administered twice within each condition week—at the start-of-week (t1) and end-of-week (t2) assessments—enabling the POMS timing and carryover analyses. The MEQ and screening questionnaires were administered at intake (baseline).

### Data processing

#### Fitbit sleep data

Sleep was recorded using a wrist-worn Fitbit Charge HR. Parameters—total sleep duration, sleep efficiency, sleep onset, wake time, time in bed, and sleep midpoint—were derived from Fitbit’s automatic sleep-detection output. Nights with incomplete recordings (<3 hours total sleep) were excluded prior to computing weekly summaries. Weekly means were calculated for each condition, and within-person weekly standard deviations were computed for variability measures, including the primary sleep-regularity outcome. Participants with no valid nights in a condition week were excluded from Fitbit-based analyses. Paired weekly contrasts required data in both weeks, whereas night-level sensitivity models used all available valid nights.

#### Daily diary data

Diary-derived variables included bedtime, wake time, time in bed, sleep onset latency, subjective sleep quality, naps, caffeine intake, and exercise duration. Unless specified, variables were summarised at the weekly condition level for crossover analyses.

#### Cognitive task data

For reaction-time tasks, trials <150 ms or >2000 ms were excluded prior to averaging, and error trials were omitted from reaction-time analyses. Stroop congruent reaction time was log-transformed. Verbal span was computed as the mean of digit, letter, and word spans; Corsi span was analysed separately for forward and backward conditions.

### Statistical analysis

#### Crossover models

All weekly and endpoint outcomes were analysed using linear mixed-effects models fitted by restricted maximum likelihood (REML):

`outcome ∼ condition + period + sequence + (1 | participant)`

Condition (BLT vs control), period (Week 1 vs Week 2), and sequence (BLT→control vs control→BLT) were included as fixed effects, with participant as a random intercept. The condition effect estimates the within-participant BLT–control difference, adjusted for period and sequence. Models were fitted using lme4 and lmerTest, with Satterthwaite degrees of freedom and the bobyqa optimiser. Condition coefficients were coded as BLT minus control (positive = higher under BLT).

#### KSS and SSS diurnal profile models

For repeated KSS and SSS ratings, models included time of day and the condition × time interaction. Primary analyses used four daytime assessments (12:00, 15:00, 18:00, 21:00), with time coded as an ordered numeric predictor (0–3) to test linear diurnal change. Follow-up contrasts were estimated at each time point. Bedtime ratings were excluded from the primary model.

#### Multiple-comparison correction

Primary outcomes were adjusted using the Holm sequential Bonferroni procedure. Secondary and exploratory analyses were reported with uncorrected p-values.

#### Mechanistic and sensitivity analyses

Exploratory analyses tested whether BLT effects accumulated across days; altered coupling between prior-night sleep deviation and next-day sleepiness; shifted sleep midpoint, MEQ alignment, or social jetlag; and whether sleep regularity mediated the KSS 12:00 effect. Sensitivity analyses addressed POMS timing and carryover, Fitbit missingness and a diary-based bedtime-variability proxy, and baseline-adjusted cognitive models.

#### Software

Analyses were conducted in R (version 4.5.1) using tidyverse, lme4, lmerTest, emmeans, effectsize, and pwr. Spectral calculations were based on measured spectra and CIE S 026 α-opic metrology.

### Qualitative analysis

After study completion, all 33 participants completed brief semi-structured interviews in Mandarin Chinese. The interviews compared BLT and control across sleep, alertness, mood, cognition, and acceptability.

Responses were analysed using directed framework coding derived from trial domains and extended to include acceptability, side effects, and contextual factors. Computer-assisted coding was applied to all 165 response units and verified manually by the first author. Each response was assigned a directional code (improvement, no difference, worsening, mixed, uncertain/confounded, or none). Selected quotations were translated into English.

### Sample size and power

A paired-design calculation (two-tailed α = .05) indicated that 33 participants provided ∼80% power to detect a medium within-person effect (Cohen’s dz = 0.50), but not small effects.

## Results

### Participants and completion

Thirty-three participants completed both conditions of the crossover trial (17 BLT-first, 16 Control-first). Table 1 summarises baseline characteristics. Mean age was 24.7 ± 2.4 years (range 18–30); 24 participants were female (72.7%). MEQ scores indicated predominantly intermediate to moderate-evening chronotype (mean 42.1 ± 8.1; range 32–60: 2 moderate morning, 13 intermediate, 18 moderate evening). There were no dropouts after enrolment. All participants completed all seven scheduled morning BLT sessions (100% session adherence). No serious adverse events were reported. In the post-trial interviews, 12 of 33 participants (36%) reported negative or mixed experiences with BLT acceptability—most commonly minor, generally transient eye discomfort or glare (typically resolving after three to four days), with less frequent dizziness or nausea, sweating during sessions, and, in one participant, increased impatience during the BLT week. Four participants had no usable Fitbit data, reducing the Fitbit subsample to N = 29; one additional participant contributed Fitbit data in the BLT week only and so could not enter the within-person comparison, leaving paired Fitbit analyses based on 28 participants (27 for sleep-architecture variability). The four participants with no usable Fitbit data were slightly older than participants with at least some usable Fitbit data (mean 26.8 vs. 24.4 years; p = .039), but there was no clear evidence of systematic differences on gender, BMI, MEQ, or control-week diary sleep variables (smallest other p = .091, for MEQ); the small number of these participants (n = 4) limits inference about the missingness mechanism (see Supplementary Table S1).

### Primary outcomes

#### Daytime sleepiness

The primary KSS model revealed a significant condition × time-of-day interaction (b = 0.211 per 3-hour step, SE = 0.059, p < .001, Holm p = .002), indicating that BLT produced a selective, time-dependent reduction in subjective sleepiness rather than a uniform shift across the day. The BLT benefit was concentrated in the first half of the day: at 12:00, BLT reduced KSS sleepiness relative to control by approximately 0.47 units (b = −0.47, SE = 0.11, p < .001, dz = −0.58), and a smaller reduction remained significant at 15:00 (b = −0.26, SE = 0.07, p < .001, dz = −0.14). The difference attenuated to non-significance by the evening, with no reliable effect at 18:00 (b = −0.05, p = .461) or 21:00 (b = 0.16, p = .153; Figure 2). The overall mean of the daytime KSS ratings did not differ between conditions (b = −0.1, p = .46), consistent with the finding that BLT effects on sleepiness were concentrated in the midday-to-early-afternoon period.

**Figure 2.**
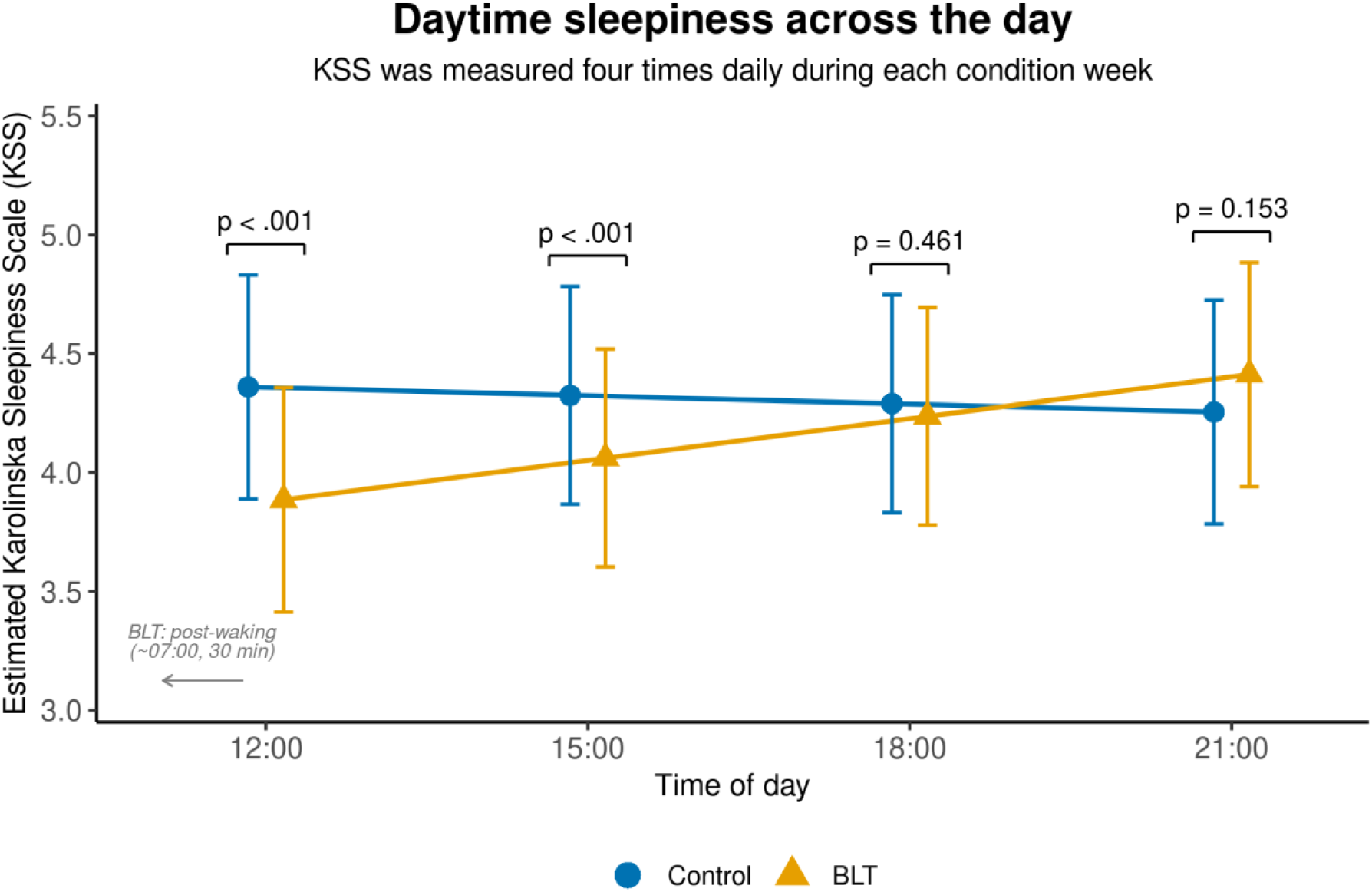
Effect of Bright Light Therapy on Daytime Sleepiness Across the Day. *Note.* Model-estimated Karolinska Sleepiness Scale (KSS) ratings at 12:00, 15:00, 18:00, and 21:00 under BLT and control. Error bars denote 95% confidence intervals. BLT = bright light therapy; KSS = Karolinska Sleepiness Scale.

#### Sleep regularity

In the Fitbit subsample, BLT significantly reduced night-to-night variability in sleep duration (SD of total sleep duration: b = −25.98 min, SE = 9.57, p = .011, Holm p = .046, dz = −0.52, N = 28; Figure 3). A sensitivity analysis using a diary-based bedtime-variability proxy in the full sample (N = 33) did not replicate this finding (b = −3.09, p = .665), indicating that the sleep-regularity result should be interpreted as applying to the Fitbit-wearable subsample rather than generalised to the full cohort.

**Figure 3.**
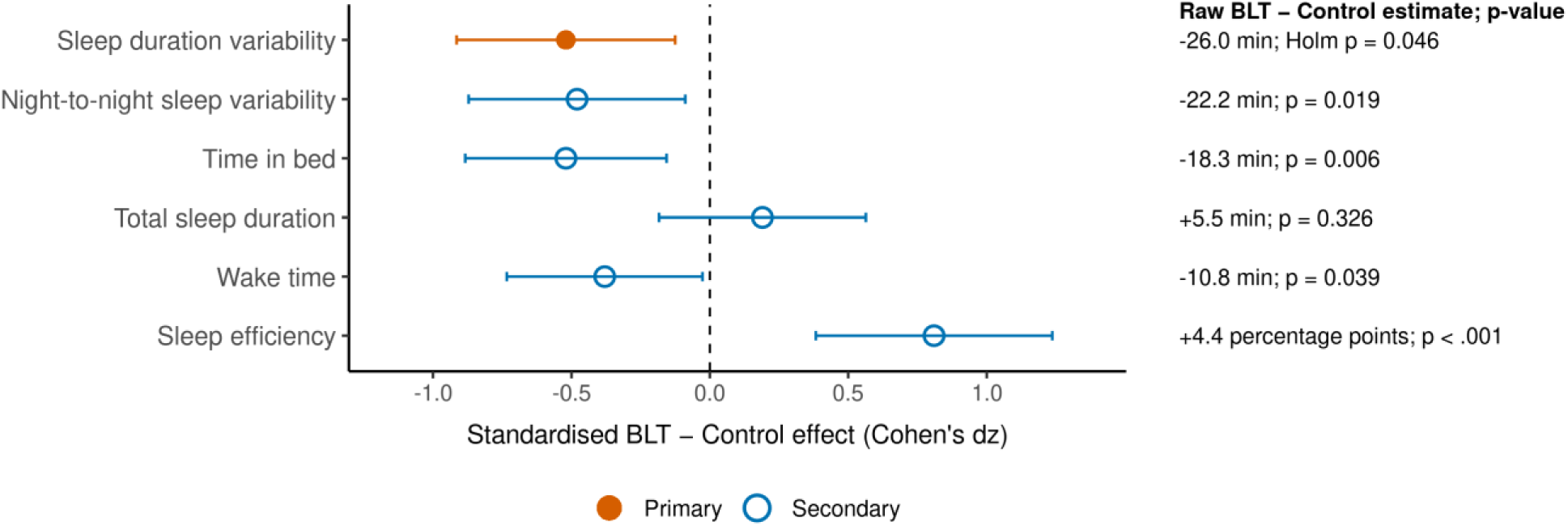
Effects of Bright Light Therapy on Fitbit-Derived Sleep Outcomes. *Note.* Forest plot of BLT-versus-control crossover estimates with 95% confidence intervals, based on the Fitbit wearable subsample with complete two-condition data (n = 28; n = 27 for sleep-architecture variability). BLT = bright light therapy; RMSSD = root mean square of successive differences.

#### Positive affect

PANAS Positive Affect was significantly higher under BLT than control (b = 2.13, SE = 0.81, p = .013, Holm p = .046, dz = 0.41), representing a medium-sized improvement in positive emotional experience associated with the BLT condition.

#### Cognitive outcomes (primary)

Neither prespecified primary cognitive outcome showed a reliable BLT-related improvement. Stroop log-transformed RT was numerically shorter under BLT but did not reach significance (b = −0.033, SE = 0.023, p = .160, Holm p = .321, dz = −0.21). Verbal span showed no meaningful difference (b = 0.061, SE = 0.113, p = .594, Holm p = .594, dz = 0.07). Full primary results are presented in Table 2.

**Table 2.** Primary Confirmatory Outcomes: Bright Light Therapy Versus Control Crossover Effects.

| Outcome | N | b (SE) | p (raw) | p (Holm) | dz |
| --- | --- | --- | --- | --- | --- |
| Daytime sleepiness: KSS condition × time-of-day interaction, per 3-h step | 33 | 0.211 (0.059) | < .001 | .002 | — |
| Sleep regularity: SD of total sleep duration, min (Fitbit) | 28 | −25.98 (9.57) | .011 | .046 | −0.52 |
| Positive affect: PANAS Positive Affect | 33 | 2.13 (0.81) | .013 | .046 | 0.41 |
| Cognition: Stroop log-transformed RT, congruent | 33 | −0.033 (0.023) | .160 | .321 | −0.21 |
| Memory: Verbal span mean | 33 | 0.061 (0.113) | .594 | .594 | 0.07 |

*Note.* Holm correction was applied across the five primary confirmatory outcomes. Positive
coefficients ( $b$ ) indicate higher values under BLT than control; negative coefficients indicate lower values under BLT. $dz$ = Cohen's paired effect size, calculated as the mean within-person BLT – control difference divided by the SD of the difference scores. $dz$ is not reported for the KSS interaction term because it describes a profile shape rather than a single mean difference. BLT = bright light therapy; KSS = Karolinska Sleepiness Scale; PANAS = Positive and Negative Affect Schedule; RT = reaction time; SE = standard error.

### Secondary outcomes

#### Sleep efficiency, time in bed, and wake time

The strongest secondary effect in the dataset was Fitbit sleep efficiency, which increased under BLT by 4.35 percentage points (b = 4.35, SE = 1.02, p < .001, dz = 0.81; N = 28).

Correspondingly, total time in bed was shorter under BLT by approximately 18 minutes (b = −18.3 min, SE = 6.2, p = .006, dz = −0.52; N = 33), suggesting that BLT was associated with more efficient consolidation of sleep within a reduced time-in-bed window. Diary-recorded wake time was approximately 11 minutes earlier under BLT (b = −10.8 min, SE = 5.0, p = .039, dz = −0.38), consistent with earlier morning activation, although sleep midpoint and other circadian-characterisation indices were not significantly shifted. A second index of night-to-night variability, the root mean square of successive differences (RMSSD) in sleep duration, was likewise reduced under BLT (b = −22.23 min, SE = 8.94, p = .019, dz = −0.48; N = 28), converging with the primary sleep-regularity finding. Sleep onset latency, total sleep duration, and sleep midpoint did not differ reliably between conditions (all p > .32). Full secondary sleep and alertness results are presented in Table 3.

**Table 3.**
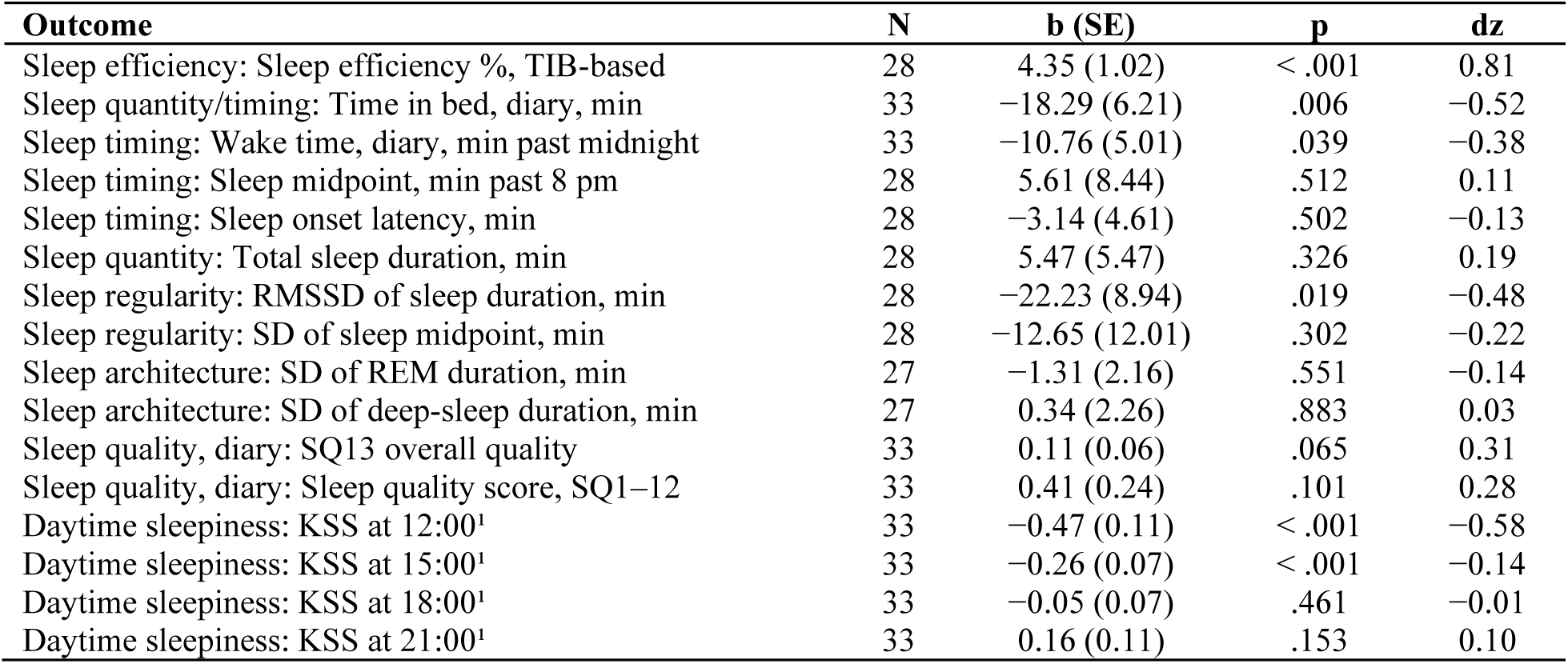
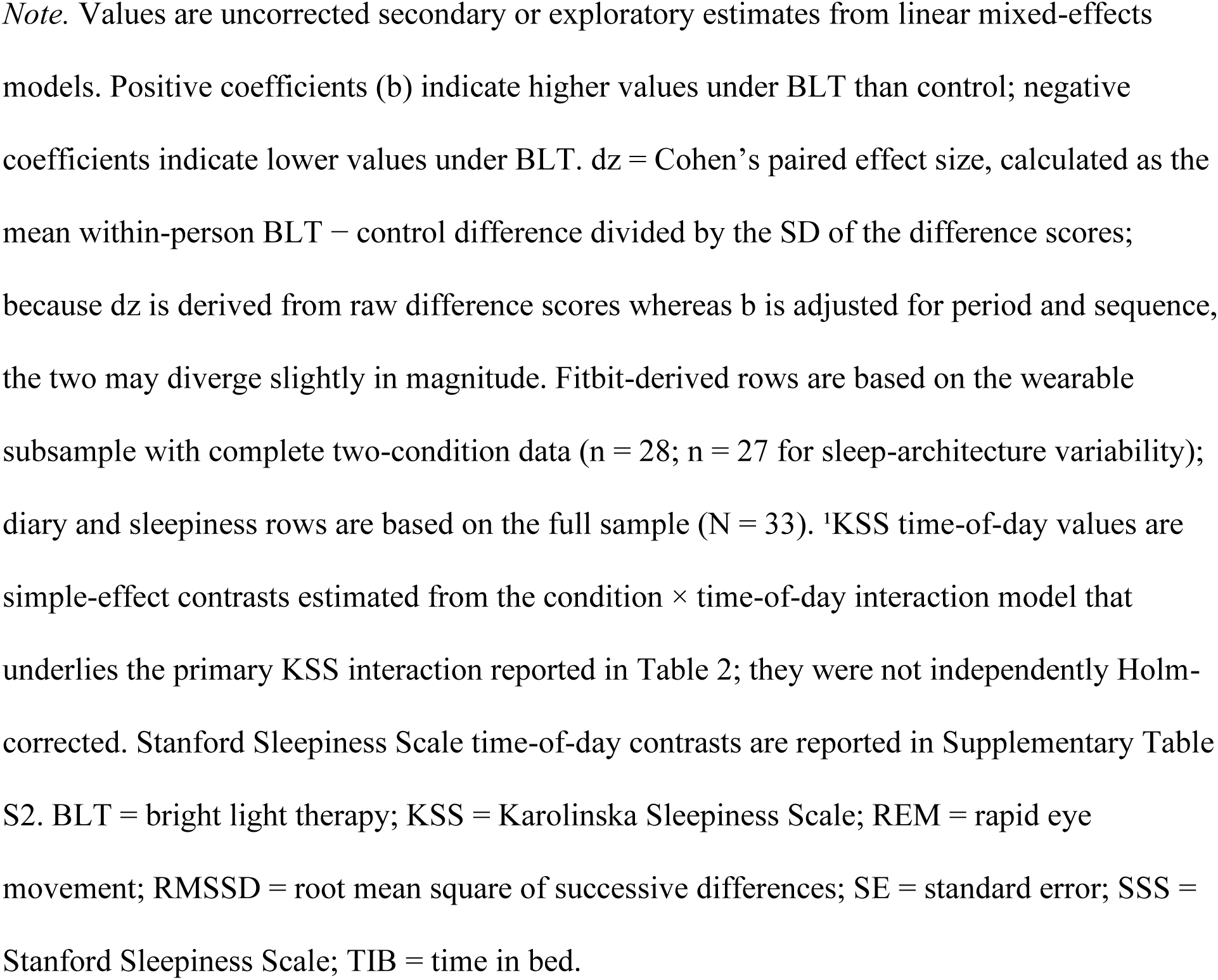
Secondary Sleep and Alertness Outcomes: Bright Light Therapy Versus Control Crossover Effects.

#### Secondary sleepiness

The Stanford Sleepiness Scale showed the same early-day pattern as the KSS, with significant reductions under BLT at 12:00 (b = −0.36, SE = 0.09, p < .001, dz = −0.47), 15:00 (b = −0.25, p < .001), and 18:00 (b = −0.14, p = .015), attenuating to no difference by 21:00. However, the SSS carryover check showed a significant sequence-dependent signal (p = .016), suggesting that SSS ratings were influenced by order of condition exposure. KSS did not show evidence of carryover (p = .401) and was therefore retained as the primary sleepiness outcome; full SSS time-of-day contrasts are reported in Supplementary Table S2 and plotted in Supplementary Figure S1.

#### Mood (secondary and exploratory)

Among POMS subscales, Total Mood Disturbance was nominally lower under BLT (b = −7.22, SE = 3.46, p = .045, dz = −0.35) and Anger-Hostility was reduced (b = −1.50, SE = 0.68, p = .034, dz = −0.38). These POMS effects are treated as exploratory because POMS was not included in the primary Holm family. Vigor-Activity showed a directionally consistent trend (b = 1.25, SE = 0.69, p = .077, dz = 0.32). The Esteem-related Affect–Positive composite did not reach significance (b = 0.61, p = .191). PANAS Negative Affect was not significantly reduced (b = −1.35, SE = 1.03, p = .198, dz = −0.23). The pattern across mood outcomes — improvement in positive affect and mood disturbance, no reliable change in negative affect — is summarised in Figure 4. Figure 4 and Supplementary Table S3 orient effect sizes so that positive values indicate benefit under BLT.

**Figure 4.**
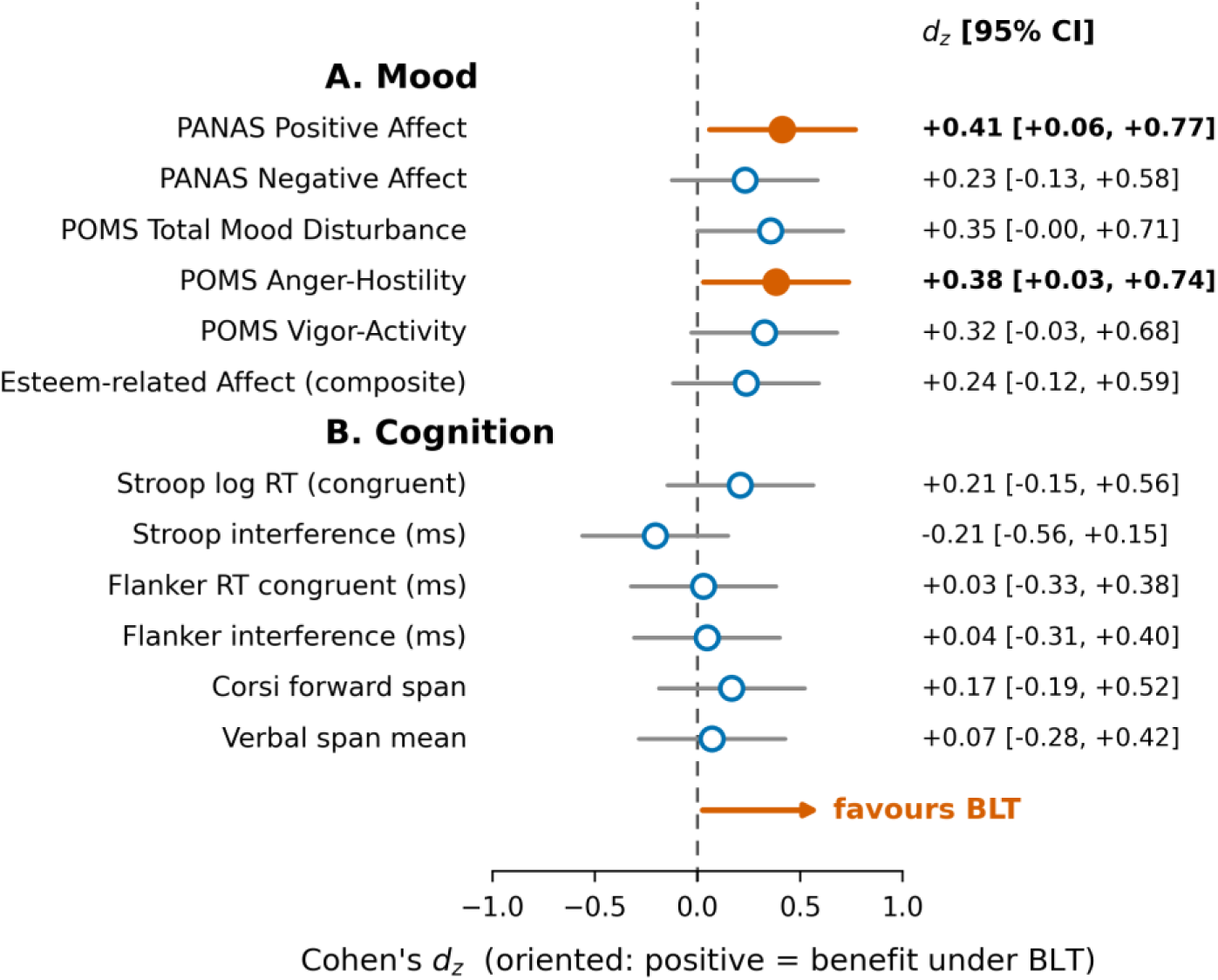
Within-Person Effects of Bright Light Therapy on Mood and Cognition. *Note.* Forest plot of Cohen’s paired effect sizes (dz) with 95% confidence intervals, oriented so that positive values indicate benefit under BLT. (A) Mood; (B) cognition. BLT = bright light therapy; PANAS = Positive and Negative Affect Schedule; POMS = Profile of Mood States.

#### Cognition (secondary)

The null pattern observed for primary cognitive outcomes extended across all secondary cognitive measures. Stroop RT (ms) and interference score, Flanker congruent RT and interference score, and Corsi forward span were all non-significant and showed small to negligible effect sizes (dz range: −0.05 to +0.21; all p > .22; see Supplementary Table S3). The consistency of near-zero cognitive effects across five independent cognitive measures strengthens the conclusion that BLT did not produce objective cognitive enhancement at this dose and duration.

### Sensitivity and validity checks

#### Period and sequence checks

All crossover models included period and sequence terms. Sequence effects were non-significant for the primary confirmatory outcomes, providing no evidence of strong order-dependent contamination in the main BLT effects (the Stanford Sleepiness Scale, a secondary outcome, showed a sequence-dependent carryover signal and is reported above). Period effects were observed for the KSS 12:00 follow-up contrast (p = .018) and PANAS Positive Affect (p = .002), indicating week-to-week shifts independent of condition. These period effects were adjusted for in the crossover models and therefore do not change the estimated BLT-versus-control contrasts, although they indicate that some outcomes were sensitive to study week. Full period, sequence, and carryover design diagnostics are reported in Supplementary Table S4.

#### POMS timing and carryover

POMS TMD and Anger-Hostility were assessed at the start (t1) and end (t2) of each condition week. Neither outcome showed a significant increase in effect size from t1 to t2 (TMD delta: b = −0.65, p = .326; Anger delta: b = −0.04, p = .899), indicating that mood effects did not accumulate progressively across the week. Carryover tests for POMS were non-significant (TMD: p = .858; Anger: p = .508), providing no evidence of sequence-dependent carryover for these mood outcomes. Full POMS timing and carryover analyses are reported in Supplementary Table S5.

#### Mechanistic and circadian checks

Daily-buildup models showed no evidence that BLT effects progressively accumulated across the intervention week for KSS at 12:00 (slope b = −0.051, p = .374), sleep duration (slope b = 3.52 min, p = .300), or sleep-duration deviation (slope b = −1.66, p = .438). Circadian-characterisation analyses also found no reliable BLT-related shifts in sleep midpoint (b = 5.6 min, p = .512), circadian alignment (b = 5.3 min, p = .535), or social jetlag (b = 5.3 min, p = .648), indicating that the observed alertness and sleep-efficiency benefits were not accompanied by a detectable shift in the timing of the sleep episode. A bootstrapped mediation analysis (1,000 draws) found no evidence that Fitbit sleep regularity mediated the BLT effect on midday sleepiness (indirect effect = −0.047, 95% bootstrap CI [−0.261, +0.056], p = .36), providing no evidence that the measured sleep-regularity effect statistically mediated the midday sleepiness effect; together, these analyses suggest that the KSS and sleep-regularity effects were not explained by a simple cumulative phase-shift or sleep-regularity mediation pathway. Daily covariates — exercise duration, outdoor time, and caffeine consumption — did not differ significantly between conditions (all p > .08) and are unlikely to account for the observed effects. Full mechanistic, circadian, and mediation results are reported in Supplementary Table S6, and daily trajectories across the intervention week are shown in Supplementary Figure S2.

### Cognitive baseline adjustment

Inclusion of pre-trial baseline scores as covariates in the Stroop and verbal span models produced estimates that were numerically identical to the unadjusted crossover models (Stroop log RT: b = −0.033, p = .160; verbal span: b = 0.061, p = .594). The cognitive null was therefore robust to individual differences in pre-trial cognitive performance.

### Explanatory interview findings

Post-trial interviews provided explanatory context for the quantitative findings (Figure 5). For sleep quality (Q1), 19 participants (57.6%) reported perceived improvement under BLT, 9 (27.3%) reported no clear difference, and 4 (12.1%) described uncertainty or acknowledged confounding factors. For sleep pattern and alertness (Q2), 16 participants (48.5%) reported improvement, most commonly easier morning awakening or greater midday alertness, while 11 (33.3%) reported no clear difference. Mood effects were less consistent (Q3): 10 participants (30.3%) reported improved mood or emotional wellbeing, 15 (45.5%) reported no clear mood difference, and 6 (18.2%) acknowledged uncertainty or contextual confounding from workload or stress. For subjective cognition (Q4), 16 participants (48.5%) reported perceived improvement in concentration or mental clarity; however, these reports should be interpreted as perceived cognitive clarity rather than objective cognitive enhancement, given the uniformly null objective findings. Additional comments on BLT experience and acceptability (Q5) highlighted tolerability and feasibility concerns: 10 responses (30.3%) described negative experiences and 2 (6.1%) described mixed experiences. Aggregated directional-code counts are reported in Supplementary Table S7, and a participant-level heatmap of individually coded responses is shown in Supplementary Figure S3.

**Figure 5.**
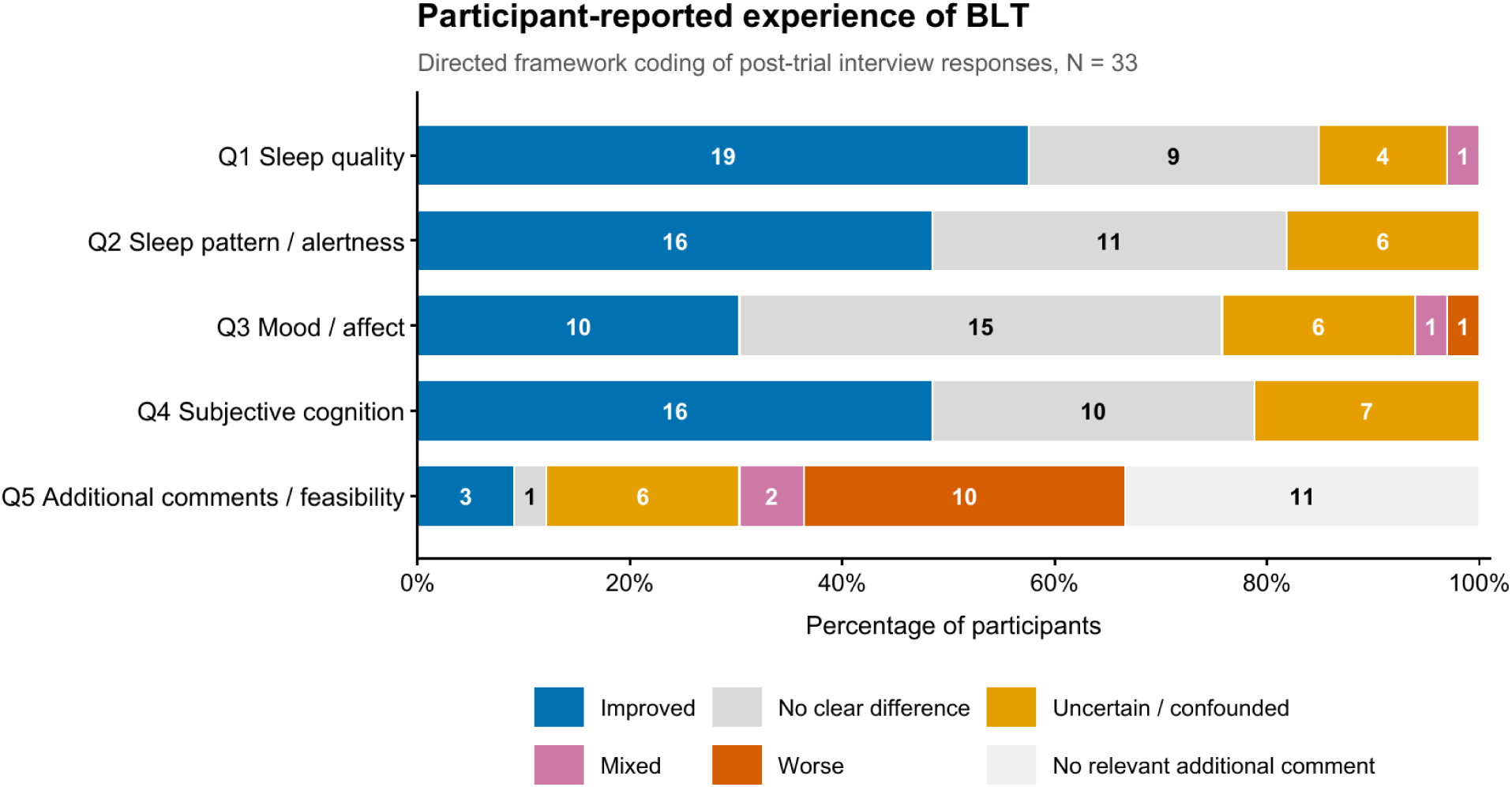
Participant-Reported Experiences of Bright Light Therapy. *Note.* Directed framework coding of post-trial semi-structured interview responses (N = 33) across five domains (Q1, sleep quality; Q2, sleep pattern and alertness; Q3, mood and affect; Q4, subjective cognition; Q5, additional comments and feasibility). Horizontal 100% stacked bars show the percentage of participants assigned to each directional code, with participant counts printed inside the bars. Codes are improved, no clear difference, uncertain/confounded, mixed, worse, and—for Q5—no relevant additional comment. Q4 reflects subjective cognitive experience rather than objective task performance, which showed no BLT-related change. BLT = bright light therapy.

The qualitative direction-code pattern converges with the quantitative results for alertness and sleep, partly converges for mood, and diverges for cognition. Selected illustrative quotations are provided below.

**Sleep quality and alertness:** “During the light therapy week I slept more deeply; without light therapy I would wake briefly during the night and had more difficulty getting up in the morning.” (w2.3, Q1)

“The routine was similar, but without light therapy I especially wanted to nap at noon; with light therapy I did not want to nap as much.” (w3.8, Q2)

“After light therapy I more easily shook off sleepiness after waking and became alert; the time to fall asleep also seemed slightly shorter.” (w3.6, Q1)

**Mood benefit and confounding:** “Emotionally I felt more relaxed; before light therapy I was more anxious, but during the light therapy week my mood felt more comfortable.” (w1.4, Q3)

“It was very difficult to separate the effect of light therapy on my mood from the much larger effect of my research tasks.” (w1.2, Q3)

**Subjective cognition:** “For attention, I could start working faster, and mental activity felt more flexible, but it did not fundamentally change my work ability.” (w4.4, Q4)

**Acceptability and side effects:** “Because the light intensity was strongest, my eyes felt uncomfortable at first and sometimes it was hard to complete the full 30 minutes.” (w2.1, Q5)

Overall, interviews suggest that BLT was experienced primarily as a sleep/alertness intervention, with supportive but less consistent affective benefits, and subjective cognitive improvements that were not confirmed by objective task performance.

## Discussion

In this randomized two-week crossover trial, one week of morning bright light therapy (BLT), compared with one week of usual-light control, produced a coherent pattern of benefits across sleep and daytime alertness in healthy university students, while leaving objective cognitive performance unchanged. The clearest effects were a time-of-day–specific reduction in subjective sleepiness largest at midday, a reduction in night-to-night variability of sleep duration on two converging indices, an increase in Fitbit-derived sleep efficiency, and a small improvement in positive affect. Objective measures of processing speed and working memory showed no reliable change.

The clearest confirmatory effect was on daytime sleepiness. BLT did not lower sleepiness uniformly across the day; instead it produced significant reductions at 12:00 and 15:00 that diminished to negligible differences by the evening. This time-dependent profile is consistent with the established alerting action of light (Cajochen, 2007) and with experimental work showing that daytime bright-light exposure reduces sleepiness and improves vigilance (Phipps-Nelson et al., 2003). It also fits with the broader, albeit heterogeneous, literature on the acute alerting effect of light, wherein higher intensity has been shown to enhance subjective alertness in many, but not all, instances (Souman et al., 2018). The concentration of the benefit in mid-to-early-afternoon is mechanistically explained. Morning light exposure, like light at any time, has an acute enhancing effect on alertness mediated via the non-image-forming (NIF) pathway (Berson et al., 2002), complemented by this time coinciding with the rising part of the circadian alerting signal. Morning exposure also falls on the phase-advancing region of the human phase response curve to light (Khalsa et al., 2003), although we did not detect any significant shift in sleep midpoint or other circadian indices. This suggests the alerting value of morning light lies in its acute NIF effect, which may counter the midday dip. That the overall daily mean of the KSS did not differ between conditions underscores that BLT reshaped the diurnal profile rather than producing a constant downward shift—an interpretation reinforced by the convergent, though carryover-affected, pattern in the Stanford Sleepiness Scale.

BLT also stabilized and consolidated sleep. Two indices of night-to-night variability in sleep duration, its standard deviation and root mean square of successive differences, both decreased. Sleep efficiency rose, accompanied by shorter time in bed and slightly earlier wake times, while total sleep duration was unchanged. Reduced sleep-duration variability is particularly relevant here because irregular sleep–wake patterns are associated with delayed circadian timing and poorer academic performance in undergraduates (Phillips et al., 2017), and with greater social jetlag (Wittmann et al., 2006). A plausible mechanism here is that a fixed morning light regularizes wake time without producing detectable shifts in the sleep episode.

This is consistent with the absence of significant differences in sleep midpoint and circadian indices between our experimental conditions. Two caveats temper our findings: the regularity and efficiency results derive from consumer actigraphy (Fitbit), which is not polysomnography (Haghayegh et al., 2019), and the sleep-regularity effect did not replicate in the diary-based proxy. Second, because total sleep duration did not change, BLT appears to have improved the patterning and efficiency of sleep rather than its quantity.

The effect of BLT on mood, while small, was in the expected direction. Positive affect improved, but negative affect was unchanged. BLT nominally lowered total mood disturbance and scores on the anger–hostility dimension. The direction is consistent with the well-documented antidepressant action of bright light, which has shown effect sizes comparable to pharmacotherapy in seasonal and non-seasonal depression (Golden et al., 2005; Lam et al., 2016). Our effects were small, as expected in healthy students without clinically significant mood symptoms, leaving little room for significant improvement. The improvement was concentrated in positive rather than in negative affect, and may indicate that in non-clinical populations morning light acts more on energy and engagement than on distress.

The light dose was comparable to standard clinical BLT protocols and carefully characterized. The light stimulus we used was designed to match the intensity and duration of standard bright-light protocols used in affective and circadian disorders (Golden et al., 2005; Lam et al., 2016), providing strong stimulation of the NIF pathway (Berson et al., 2002; Lucas et al., 2014). Reporting the stimulus in melanopic units, as recommended for contemporary light research and lighting guidance (Brown et al., 2022), should aid replication and dose–response comparisons with literature that has historically described light in photopic lux alone.

In contrast to sleep, alertness, and mood, BLT produced no reliable change in cognitive functioning. Neither processing speed (Stroop) nor working memory (verbal span) improved, and this pattern extended to all secondary cognitive measures. We view this finding as informative rather than merely absent. Light can acutely modulate brain networks supporting attention and executive function (Vandewalle et al., 2009), but such effects are typically state-dependent and most evident under conditions of sleep deprivation. Three design features of our protocol provide a likely explanation of our null results in the cognitive measures. First, the cognitive battery was administered once per condition week rather than during peak light exposure. Second, the dose and duration may not have been sufficiently powerful to translate into measurable performance gains in healthy students. Lastly, the study was powered for medium rather than small within-person effects. Notably, many participants reported improved concentration and mental clarity under BLT, a dissociation worth highlighting, since conflating subjective alertness with measured performance could lead to overstating the cognitive benefits of light exposure.

Our qualitative findings strengthen the quantitative results. Participants most often reported perceived improvements in sleep quality and morning alertness, converging with the KSS and Fitbit-derived sleep-regularity effects, whereas their reports about mood and cognition were more mixed, consistent with our results on mood and cognition. These convergences and divergences illustrate the value of a mixed-methods design for distinguishing reliable benefits from subjective perceptions. What a person perceives may or may not match objective outcomes, a pattern commonly seen in the sleep and circadian literature. From a translational standpoint, the intervention was brief and self-administered. Critically, diary reporting was complete and no serious adverse events occurred; only 12 of 33 participants (36%) reported negative or mixed acceptability experiences. The latter were most commonly minor and transient eye discomfort or glare associated with the light stimulus. For a student population in whom irregular sleep is academically consequential (Phillips et al., 2017), a non-clinical morning light routine is an attractive, scalable target for campus well-being programs, which is also relevant to the new daylight exposure recommendations (Brown et al., 2022).

The triangulation of wearable, diary, questionnaire, cognitive, and interview data, together with detailed melanopic characterization of the light stimulus, is particularly relevant to modern lifestyles. However, a few limitations of the study serve as caveats, tempering the interpretation of our results. Because the two conditions ran in consecutive weeks without a washout, period effects could not be fully excluded. Also, the midday KSS and positive affect outcomes showed week effects independent of condition, suggesting possible adaptation or temporal-context influences. The trial was open-label without a sham control, so expectancy effects cannot be ruled out for subjective outcomes. Sleep metrics relied on consumer actigraphy rather than polysomnography, and four participants lacked usable Fitbit data, reducing paired analyses to 28 participants. Adverse events were captured through daily contact and diary records rather than a structured checklist, which may have under-recorded minor symptoms. The sample comprised healthy young adults of predominantly evening and intermediate chronotype in shared student housing, limiting generalizability. Finally, the modest sample provided adequate power only for medium within-person effects, so small but genuine effects, particularly on cognition, may have gone undetected.

In conclusion, one week of self-administered morning BLT does improve daytime alertness, most strongly at midday, regularizes sleep, and improves positive affect. Crucially, morning light is a good, low-burden strategy for students to stay alert, feel positive, and sleep well. Future studies using controlled trials with washout periods, objective circadian phase markers, research-grade sleep measurement, and standardized melanopic dosing are needed to extend our findings.

## Declarations

## Ethics approval and consent to participate

The study protocol was approved by the Institutional Review Board of The Hong Kong Polytechnic University (reference number HSEARS20240430005) and was conducted in accordance with the Declaration of Helsinki. All participants provided written informed consent.

## Data availability

The data that support the findings of this study are available from the corresponding author upon reasonable request.

## Trial registration

This study was preregistered at ClinicalTrials.gov (identifier NCT07685262; https://clinicaltrials.gov/study/NCT07685262).

## Data Availability

All data produced in the present study are available upon reasonable request to the authors

