## Supplementary Material (Tables S1-S8 and Figures S1-S3) for "Effects of Morning Bright Light Therapy on Sleep, Alertness, Mood, and Cognition in Healthy University Students: A Randomized Crossover Trial"

This file contains Supplementary Tables S1–S7 and Supplementary Figures S1–S3. Unless otherwise noted, estimates are from linear mixed-effects models of the form outcome ~ condition + period + sequence + (1 | participant), with positive coefficients (b) indicating higher values under bright light therapy (BLT) than control. dz = Cohen's paired effect size (mean within-person BLT − control difference / SD of the difference scores).

### Supplementary Table S1

*Fitbit Missingness and Diary-Proxy Sensitivity*

*Panel A. Participants with versus without usable Fitbit data*

| Variable | Fitbit available (n = 29) | No Fitbit (n = 4) | p |
|---|---|---|---|
| Gender (% female) | 72% | 75% | 1.00 |
| Age (years) | 24.4 (2.4) | 26.8 (1.5) | .039 |
| BMI (kg/m²) | 20.8 (3.5) | 21.5 (2.2) | .599 |
| MEQ total score | 42.7 (8.4) | 37.8 (4.0) | .091 |
| Time in bed — Control (min) | 501.6 (59.1) | 504.1 (21.7) | .873 |
| Wake time — Control (min past midnight) | 564.6 (69.4) | 566.2 (51.4) | .957 |
| Bedtime — Control (min past 8 PM) | 308.0 (72.3) | 302.1 (56.3) | .858 |
| Tiredness on waking — Control | −0.1 (0.9) | −0.4 (0.4) | .269 |

*Panel B. Sleep-regularity result versus full-sample diary proxy*

| Measure | b (SE) | p | N |
|---|---|---|---|
| Sleep duration SD — Fitbit (primary) | −25.98 (9.57) | .011 | 28 |
| Bedtime SD — diary proxy (full sample) | −3.09 (7.08) | .665 | 33 |

*Note.* Panel A compares the four participants without usable Fitbit data to participants with at least some usable Fitbit data (Welch t-tests; Fisher exact for gender); group differences were non-significant except for age. Panel B shows that the primary Fitbit sleep-duration-variability effect (N = 28 complete pairs) did not replicate in a full-sample diary-based bedtime-variability proxy, indicating that the regularity result should be interpreted as applying to the wearable subsample. BMI = body mass index; MEQ = Morningness–Eveningness Questionnaire; SD = standard deviation; SE = standard error.

**Supplementary Table S2**

*Stanford Sleepiness Scale Time-of-Day Contrasts: Bright Light Therapy Versus Control*

| Time of day | b (SE) | p | dz |
|---|---|---|---|
| 12:00 | −0.36 (0.09) | < .001 | −0.47 |
| 15:00 | −0.25 (0.06) | < .001 | −0.18 |
| 18:00 | −0.14 (0.06) | .015 | −0.31 |
| 21:00 | −0.03 (0.09) | .715 | −0.03 |

*Note.* Simple-effect contrasts (BLT − control) estimated from a linear mixed-effects model with a condition × time-of-day interaction, period and sequence covariates, and a random intercept for participant (full sample, N = 33). Negative coefficients (b) indicate lower sleepiness under BLT. Significant early-day reductions were observed at 12:00, 15:00, and 18:00, attenuating by 21:00. SSS ratings showed a significant sequence-dependent carryover signal ($p = .016$); the Karolinska Sleepiness Scale was therefore retained as the primary sleepiness measure. SE = standard error; SSS = Stanford Sleepiness Scale.

**Supplementary Table S3**

*Mood and Cognitive Outcomes: Effect Sizes for Primary and Secondary Measures*

*Panel A. Mood*

| Outcome | dz [95% CI] | p |
|---|---|---|
| PANAS Positive Affect | +0.41 [+0.06, +0.77] | .013 |
| PANAS Negative Affect | +0.23 [−0.13, +0.58] | .198 |
| POMS Total Mood Disturbance | +0.35 [0.00, +0.71] | .045 |
| POMS Anger-Hostility | +0.38 [+0.03, +0.74] | .034 |
| POMS Vigor-Activity | +0.32 [−0.03, +0.68] | .077 |
| Esteem-related Affect (composite) | +0.24 [−0.12, +0.59] | .191 |

*Panel B. Cognition*

| Outcome | dz [95% CI] | p |
|---|---|---|
| Stroop log RT (congruent) | +0.21 [−0.15, +0.56] | .160 |
| Stroop interference (ms) | −0.21 [−0.56, +0.15] | .253 |
| Flanker RT congruent (ms) | +0.03 [−0.33, +0.38] | .833 |
| Flanker interference (ms) | +0.04 [−0.31, +0.40] | .801 |
| Corsi forward span | +0.17 [−0.19, +0.52] | .257 |
| Verbal span mean | +0.07 [−0.28, +0.42] | .594 |

*Note.* N = 33. Effect sizes are oriented so that positive dz indicates benefit under BLT. PANAS Positive Affect, Stroop log RT, and verbal span were included in the primary Holm-corrected outcome family; all other outcomes in this table were secondary or exploratory. All p-values shown in this table are uncorrected; Holm-adjusted p-values for the primary-family outcomes are reported in main-text Table 2. The Esteem-related Affect composite is a POMS-derived positive-affect index. PANAS = Positive and Negative Affect Schedule; POMS = Profile of Mood States; RT = reaction time.

**Supplementary Table S4**

*Period, Sequence, and Carryover Checks (Design Diagnostics)*

| Outcome | Period b | Period p | Sequence b | Sequence p |
|---|---|---|---|---|
| Sleep duration SD | +13.76 | .162 | −2.48 | .834 |
| Total sleep duration | +6.14 | .271 | +3.90 | .842 |
| Sleep efficiency | +0.24 | .818 | +2.67 | .332 |
| Time in bed | +4.50 | .474 | −10.35 | .569 |
| KSS at 12:00 (follow-up contrast) | −0.36 | .018 | −0.32 | .560 |
| PANAS Positive Affect | −2.75 | .002 | −2.17 | .299 |
| POMS Total Mood Disturbance | −4.34 | .219 | +1.44 | .795 |

*Note.* Period (week) and sequence effects from the crossover models; coefficients (b) are unstandardized, in each outcome's original units. Sequence effects were non-significant for all listed outcomes. Period effects were present for the KSS 12:00 follow-up contrast and PANAS Positive Affect and were adjusted for in all models. The Stanford Sleepiness Scale (a secondary outcome) showed a significant sequence-dependent carryover signal and was therefore not retained as a primary sleepiness measure. KSS = Karolinska Sleepiness Scale; PANAS = Positive and Negative Affect Schedule; SD = standard deviation.

**Supplementary Table S5**

*POMS Timing and Carryover Analyses*

| Outcome | Contrast | b (SE) | p |
|---|---|---|---|
| POMS Total Mood Disturbance | BLT − Control at t1 | −6.57 (3.37) | .060 |
| POMS Total Mood Disturbance | BLT − Control at t2 | −7.22 (3.46) | .045 |
| POMS Total Mood Disturbance | Change (t2 − t1) | −0.65 (0.65) | .326 |
| POMS Anger-Hostility | BLT − Control at t1 | −1.47 (0.70) | .046 |
| POMS Anger-Hostility | BLT − Control at t2 | −1.50 (0.68) | .034 |
| POMS Anger-Hostility | Change (t2 − t1) | −0.04 (0.30) | .899 |

*Note*. POMS Total Mood Disturbance and Anger-Hostility were assessed at the start (t1) and end (t2) of each condition week. The non-significant Change (t2 − t1) contrasts indicate that mood effects did not accumulate across the week. Carryover (sequence) tests were non-significant (POMS Total Mood Disturbance, $p = .858$; POMS Anger-Hostility, $p = .508$), providing no evidence of sequence-dependent carryover for these mood outcomes. POMS = Profile of Mood States; SE = standard error.

**Supplementary Table S6**

*Mechanistic, Circadian, and Mediation Analyses*

*Panel A. Daily build-up across the intervention week*

| Outcome | Day-1 b (SE) | Slope b (SE) | Slope p |
|---|---|---|---|
| KSS at 12:00 | −0.38 (0.20) | −0.05 (0.06) | .374 |
| Sleep duration (min) | −4.49 (12.01) | +3.52 (3.39) | .300 |
| Absolute sleep-duration deviation from personal mean (min) | −7.27 (7.51) | −1.66 (2.13) | .438 |

*Panel B. Circadian characterisation (BLT − Control)*

| Outcome | b (SE) | p |
|---|---|---|
| Sleep midpoint, weekly mean (min past 8 PM) | +5.61 (8.44) | .512 |
| Circadian alignment (actual − predicted, min) | +5.30 (8.42) | .535 |
| Social jetlag \|weekend − weekday midpoint\| (min) | +5.33 (11.53) | .648 |
| Sleep midpoint daily-trajectory slope (min/day) | −3.22 | .395 |

*Note.* Panel A: daily-build-up models tested whether BLT effects accumulated across the 7-day condition week; all slopes were non-significant, providing no evidence of progressive cumulative build-up across the intervention week. Panel B: BLT did not significantly shift the timing of the sleep episode on any circadian index. A bootstrapped mediation analysis (1,000 resamples; N = 28) found no evidence that Fitbit sleep-duration regularity mediated the BLT effect on midday sleepiness (indirect effect = −0.047, 95% bootstrap CI [−0.261, +0.056], p = .36). Longitudinal night-level models (Panel A) and circadian-trajectory models (Panel B) use all available nights from the 29-participant Fitbit subsample, whereas paired weekly contrasts (Tables 2–3) require both conditions and are based on 28 participants. KSS = Karolinska Sleepiness Scale; SE = standard error.

**Supplementary Table S7**

*Qualitative Interview Coding Counts*

| Interview domain | Improved | No clear diff. | Uncertain / confounded | Mixed | Worse | No relevant comment |
|---|---|---|---|---|---|---|
| Q1 Sleep quality | 19 | 9 | 4 | 1 | 0 | 0 |
| Q2 Sleep pattern / alertness | 16 | 11 | 6 | 0 | 0 | 0 |
| Q3 Mood / affect | 10 | 15 | 6 | 1 | 1 | 0 |
| Q4 Subjective cognition | 16 | 10 | 7 | 0 | 0 | 0 |
| Q5 Additional comments / feasibility | 3 | 1 | 6 | 2 | 10 | 11 |

*Note.* Directed framework coding of post-trial semi-structured interview responses (N = 33); each participant was assigned one directional code per domain (counts sum to 33 per row). Corresponding percentages are shown in main-text Figure 5. Q4 reflects subjective cognitive experience rather than objective task performance, which showed no BLT-related change.

**Supplementary Figure S1**

*Effect of Bright Light Therapy on Stanford Sleepiness Scale Ratings Across the Day*

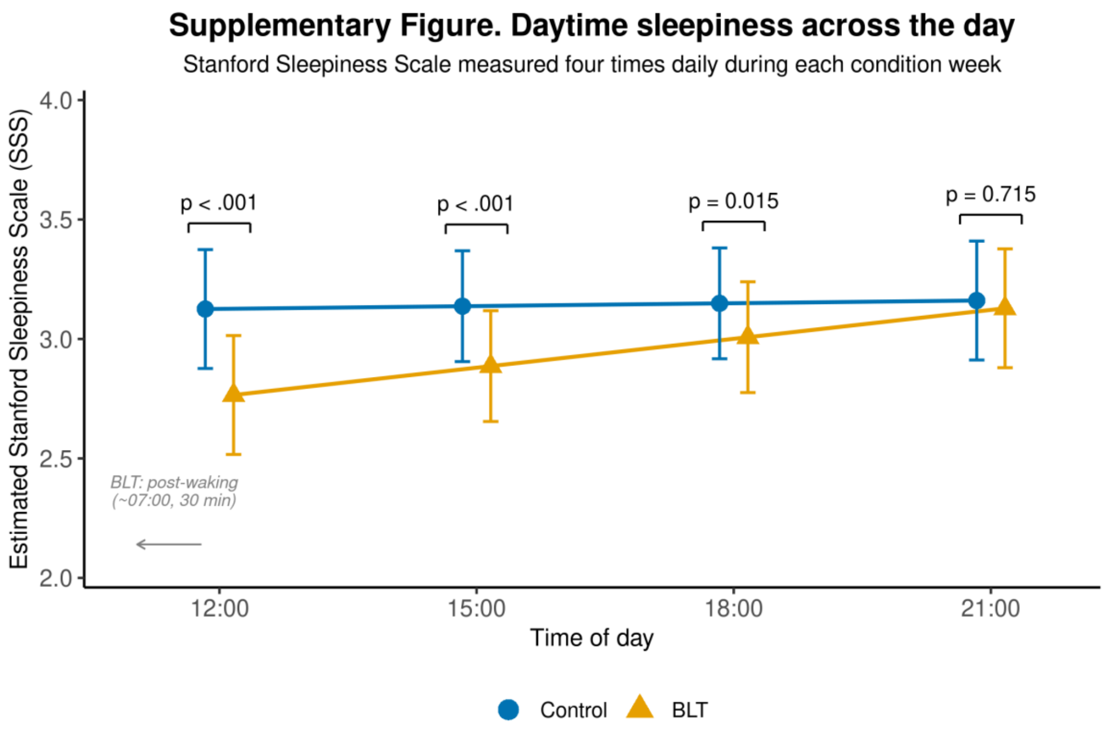


*Note.* Model-estimated Stanford Sleepiness Scale (SSS) ratings at 12:00, 15:00, 18:00, and 21:00 under bright light therapy (BLT) and control (full sample, N = 33). The SSS showed an early-day pattern consistent with the Karolinska Sleepiness Scale but a significant sequence-dependent (carryover) signal; SSS was therefore treated as secondary. Error bars denote 95% confidence intervals. Contrasts are tabulated in Supplementary Table S2. BLT = bright light therapy; SSS = Stanford Sleepiness Scale.

**Supplementary Figure S2**

*Daily Trajectories Across the Intervention Week: No Cumulative Build-Up*

(A) Midday sleepiness (KSS at 12:00)

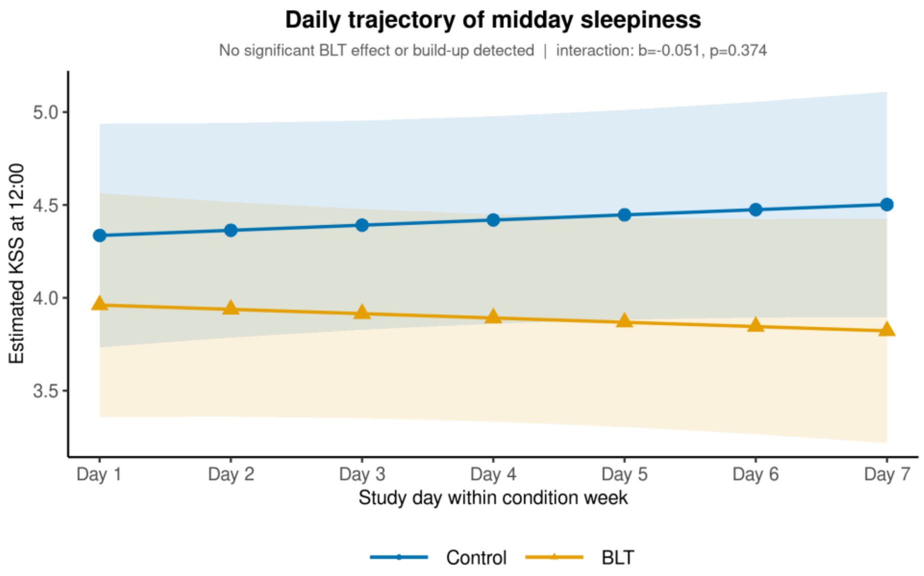


(B) Sleep duration

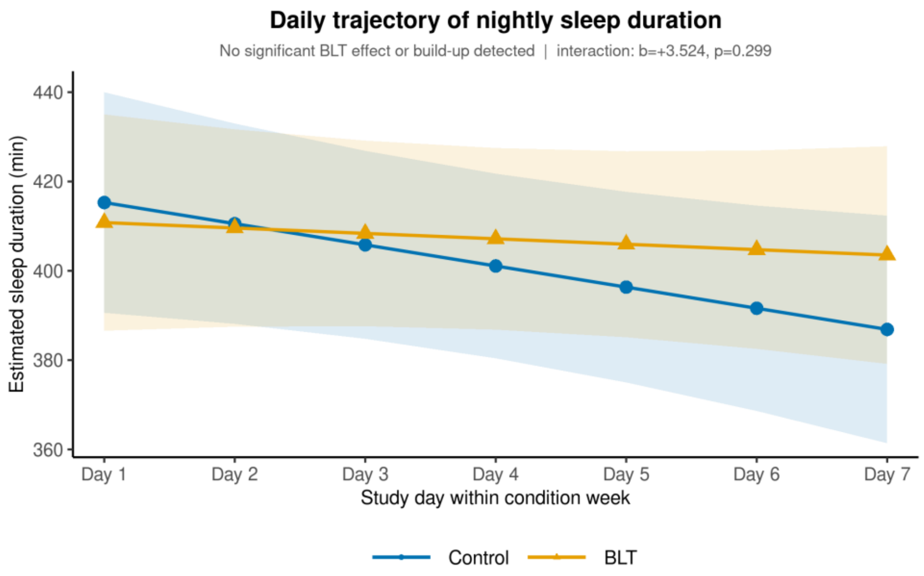


(C) Sleep-duration deviation from personal mean

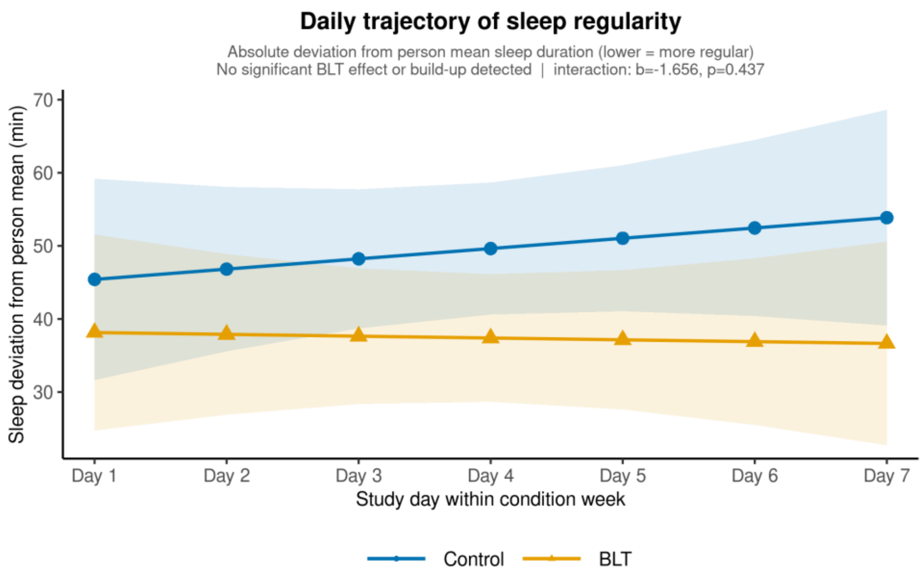


*Note.* Model-estimated daily trajectories across the 7-day condition week for (A) Karolinska Sleepiness Scale at 12:00, (B) Fitbit sleep duration, and (C) absolute deviation of sleep duration from each participant's mean, under bright light therapy (BLT) and control. None of the condition × study-day interactions was significant (all $p > .29$; see Supplementary Table S6), providing no evidence of progressive cumulative build-up across the intervention week. Shaded bands denote 95% confidence intervals. BLT = bright light therapy; KSS = Karolinska Sleepiness Scale.

**Supplementary Figure S3**

*Qualitative direction-code summary of participant-reported bright light therapy experience.*

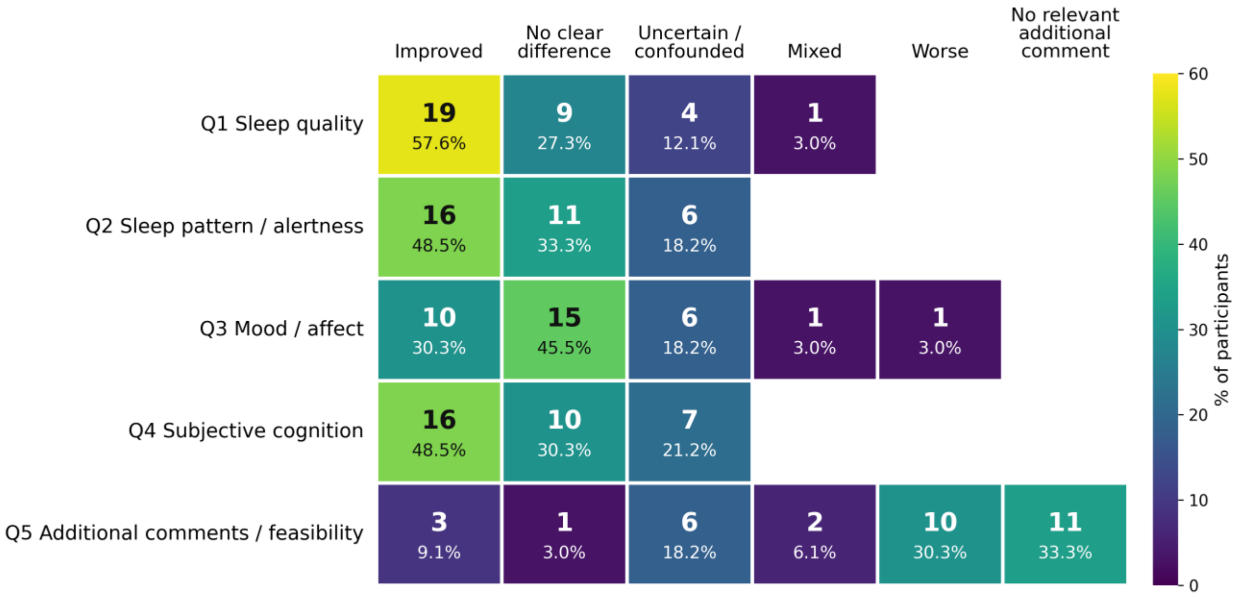


*Note.* *Directed framework coding of post-trial semi-structured interview responses (N = 33) across five interview domains (Q1–Q5: sleep quality; sleep pattern / alertness; mood / affect; subjective cognition; and overall comments / feasibility). Each cell shows the number of participants (n) and the corresponding percentage assigned to each response direction within a domain (counts sum to 33 per row); colour intensity encodes the percentage. Q4 reflects subjective cognitive experience rather than objective task performance, which showed no BLT-related change. BLT, bright light therapy.*
